# Robot-Assisted Transcranial Magnetic Stimulation Guided by Cortical Topography

**DOI:** 10.64898/2026.10.25.26366449

**Authors:** Jiaming Zhang, Yihao Liu, Jing Tian, Dale C. Roberts, Mehran Armand, Amir Kheradmand

## Abstract

Transcranial magnetic stimulation (TMS) is widely used for functional brain mapping and neuromodulation, but conventional neuronavigation systems typically rely on simplified volumetric brain models that do not fully capture subject-specific cortical geometry. We developed and evaluated a novel cortical-topography-guided framework for improving TMS planning, coil alignment, and stimulation reliability. The framework integrates an enhanced head-MRI co-registration strategy based on the iterative closest point (ICP) algorithm using a three-dimensional cortical mesh reconstructed from structural MRI. This approach was designed to improve neuronavigation by more accurately aligning the participant’s head with the MRI-derived cortical anatomy, thereby reducing co-registration error. Robot-assisted TMS (RA-TMS) was used to preserve the planned coil pose relative to cortical targets. Registration accuracy, coil alignment error, and motor-evoked potential (MEP) responses at the hand area of the primary motor cortex (M1) were evaluated and compared between RA-TMS and conventional manual TMS approach, which did not consistently preserve cortical topography-guided stimulation. ICP-based co-registration reduced the registration error between the head and MRI model from 5.83 ± 1.04 mm to 1.07 ± 0.03 mm. Compared with manual alignment, RA-TMS reduced translational error from 14.79 ± 8.03 mm to 5.22 ± 1.68 mm and rotational error from 7.01^°^ ± 2.29^°^ to 1.09^°^ ± 0.33^°^ (*p* < 0.001). RA-TMS also produced larger and more consistent MEP responses than manual TMS (0.27 ± 0.05 mV vs. 0.03 ± 0.01 mV, *p* < 0.001). Incorporating cortical topography-guided planning, combined with refined co-registration and robotic stabilization, improves coil alignment accuracy and the reliability of stimulation outcomes.

## 1. Introduction

Transcranial magnetic stimulation (TMS) provides a robust framework for probing and modulating human brain function. It is now widely used for functional brain mapping and for treating a range of neurological and psychiatric disorders (Barker et al., 1985; Cappon et al., 2022; Chaves et al., 2021; Gogulski et al., 2024; Hallett, 2000; Kahl et al., 2023; Lancaster et al., 2004; Liu et al., 2025; Otero-Millan et al., 2018; Philip et al., 2018; Richter, 2013; Ziemann et al., 2026). The effect of TMS is highly dependent on the orientation of the coil relative to the underlying brain anatomy, as it primarily targets the population of neurons that receive the maximum current induced by the magnetic field (Gomez et al., 2018; Peterchev et al., 2015; Reijonen et al., 2020; Rotenberg et al., 2014). For conventional figure-eight coils, the targeted area can extend up to a few square centimeters while accurate placement of the coil remains critical to effective stimulation (Thielscher and Kammer, 2004). Inconsistent and inaccurate TMS coil placement introduces substantial variability into stimulation outcomes, which is further compounded by physiological fluctuation in cortical excitability (Liu et al., 2025; Rodseth et al., 2017).

In current practice, TMS coil placement is typically guided by image-based neuronavigation systems that use frameless stereotaxic system to align the coil in real time with a cortical target (Comeau, 2014; Ruohonen and Karhu, 2010). This method begins by registering a series of anatomical landmarks on the participant’s head to corresponding points on their MRI. Once the head and MRI are co-registered, a trajectory marker is manually placed on the MRI to define the intended coil orientation at the target location. (Ambrosini et al., 2018; Cline et al., 2025; Matsuda et al., 2021; Noirhomme et al., 2004; Souza et al., 2018). In this approach, TMS coil is aligned tangentially to the head’s surface, with the assumption that the underlying cortex shares a similar surface topography. This assumption is based on a simplified curvilinear model of the brain that does not capture the complex geometry of the cerebral cortex. As a result, TMS coil placement fails to account for the intricate cortical topography with varying patterns of gyri and sulci at the target stimulation site (Weise et al., 2023). To address this limitation, we have developed a neuronavigation system based on fine segmentation of cortical geometry to guide TMS coil alignment. The system incorporates two key components: first, a surface-based co-registration method aligns the participant’s head with their 3D MRI model at high spatial accuracy, enabling reliable localization of cortical topography. Second, a refined coil planning algorithm computes the optimal TMS coil orientation directly from the cortical geometry at a target location. While this framework enables anatomically informed targeting of the cerebral cortex, its success depends on maintaining precise coil alignment during TMS application. Achieving such level of precision is particularly challenging with manual coil positioning, even when mechanical support is used (Zorn et al., 2012). Subtle movements of the head or the coil may disrupt proper alignment of the magnetic field with the target area, causing variability in stimulation accuracy and outcomes. Robotic systems have been introduced to address these limitations, providing improved stability and precision compared to manual coil placement, especially for long TMS sessions (Ginhoux et al., 2013; Giuffre et al., 2021; Goetz et al., 2019; Grab et al., 2018; Lancaster et al., 2004; Lebosse et al., 2007; Matsuda et al., 2021, 2024a; Noccaro et al., 2021; Pennimpede et al., 2013; Richter et al., 2013a; Shin et al., 2023; Zorn et al., 2012). However, these systems have largely been implemented without integration of cortical topography and thus do not fully exploit anatomically informed TMS coil alignment. Here, we introduce a Robot-Assisted TMS (RA-TMS) system that integrates neuronavigation guided by cortical topography for TMS coil alignment and stimulation. We first describe the key components of the system and then compare its performance with conventional manual coil positioning, in which alignment with cortical topography was not actively maintained during stimulation of the primary motor cortex (M1) in human subjects. Our findings demonstrate that TMS guided by cortical topography can reliably improve outcomes, and achieving this level of precision and accuracy requires robotic assistance, highlighting its value for both research and clinical applications.

## 2. Method

### 2.1. Coil Pose Planning

According to the principles of electromagnetic induction, magnetic flux decreases with distance, and the effect of TMS is inversely proportional to the distance from the surface of the coil to the brain cortex (Barker et al., 1985; Richter, 2013). Accurate TMS coil planning requires projecting the cortical target onto the scalp surface. When the target lies on the cortical surface, to maximize the stimulation effect, the coil should be positioned as close to the scalp as possible. Therefore, accurate coil alignment must account for both cortical topography and scalp curvature.

Here we reconstructed a 3D triangular mesh model of the brain from T1-weighted, high-resolution MRI (3T, 0.5 mm sections) using FreeSurfer (Figure 1a). This approach achieves fine brain segmentation by combining MRI models, probabilistic anatomical atlases, and topological constraints that explicitly enforce a continuous triangular cortical surface with spherical topology (Fischl, 2012). By modeling the cortex as a continuous triangular mesh, local surface orientation can be explicitly defined. The resulting surface geometry allows TMS coil trajectories to be tailored to the cortical topography by aligning the induced field perpendicular to a tangential plane at each target location (Figure 1b).

**Figure 1.**
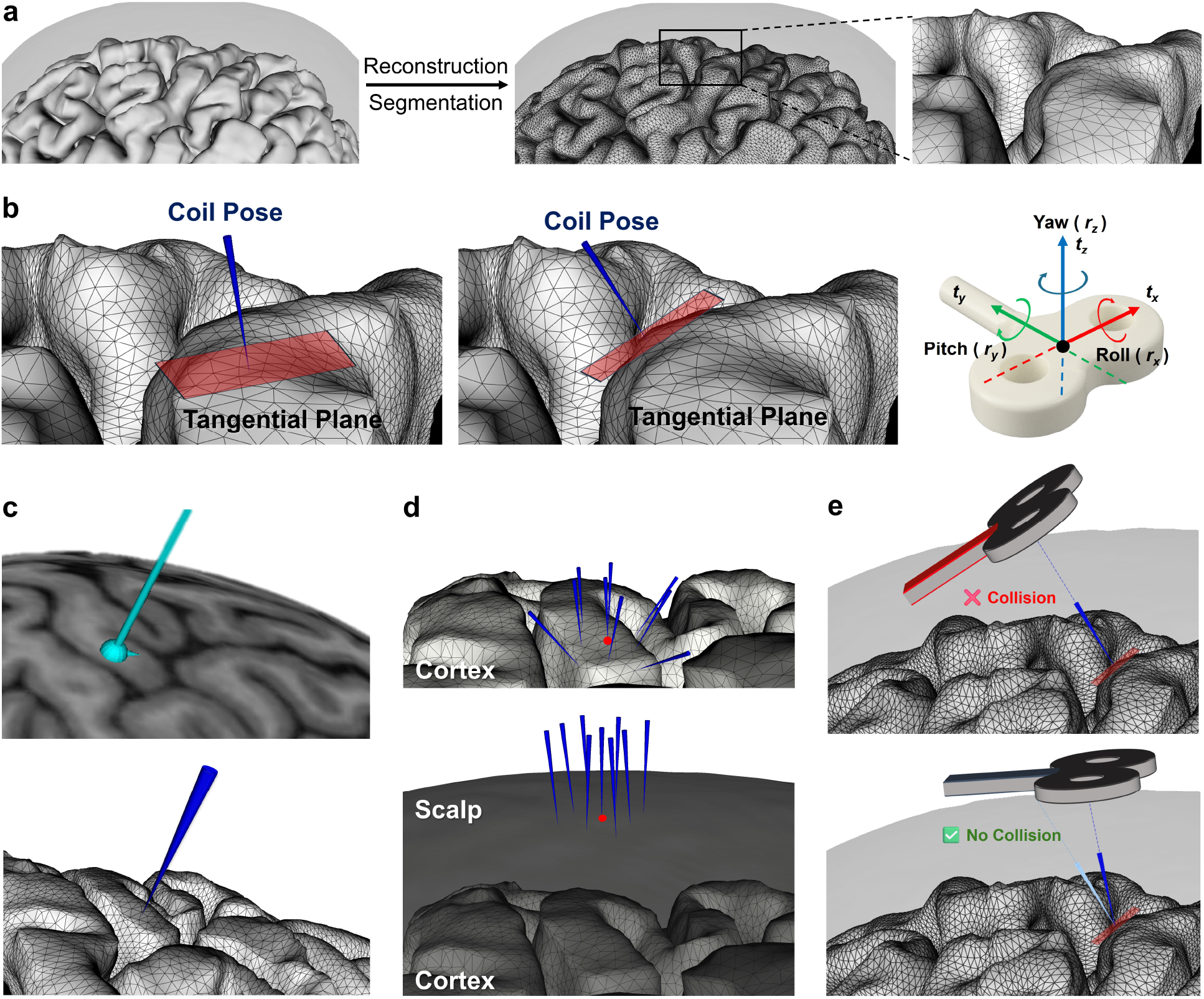
TMS guided by cortical topography. **a.** From high-resolution segmented MRI, the cortical surface is reconstructed as a triangular mesh, in which vertices are connected into triangular faces by linking neighboring points, providing detailed geometric information. Based on this representation, the TMS coil orientation can be defined as the normal vector to the local tangential plane at the target point. **b**. Two examples of coil pose computed based on selected cortical regions are shown, with the coil pose (blue cone) oriented perpendicular to the tangential plane at each target location. The TMS coil coordinate frame is shown in red, blue, and green colors, representing the X-Y-Z axis and their corresponding rotations. The coil poses shown are collinear with the Z axis of the coil coordinate frame. **c**. For comparison, a coil pose generated by a commercial neuronavigation software is shown (top), which uses a curvilinear surface model rather than cortical geometry (bottom). **d**. Comparison of a 2.5 mm-spaced grid of coil poses generated using cortical geometry (top) versus grid placement based on scalp curvature (bottom). The red point indicates the center of the grid, and the blue cones mark the coil pose trajectories for different target locations within the grid. **e**. Since the stimulation target is at the cortical surface, but the TMS coil is placed on the scalp, the planned coil pose is projected to the scalp based on the cortical target location. If the resulting orientation intersects the head, it is adjusted to the nearest scalp point from the cortical target, ensuring alignment along the shortest path between the cortex and scalp.

To incorporate cortical topography, we implemented a coil pose planning approach that directly computes coil position and orientation relative to the local cortical plane for accurate TMS delivery. In this framework, a TMS coil pose **p** is represented as a six-degree-of-freedom (6-DOF) tuple with 3D translation components (*t*_*x*_, *t* _*y*_, *t*) and 3D orientation components (*r*_*x*_, *r*_*y*_, *r*_*z*_) (Liu et al., 2025; Richter et al., 2013b) (Figure 1b). To determine the coil orientation, a point is first marked on the cortical surface (*t*_*x*_, *t*_*y*_, *t*_*z*_). A second marker defines the tail direction (Y-axis of the coil), while the local surface normal vector (*n*_*target*_) determines the Z-axis of the coil. This normal vector, which is perpendicular to the local tangential plane, is then extended outward to locate its intersection point on the scalp surface. The intersection point serves as the location of the TMS coil placement on the head, with the coil orientation defined parallel to the plane tangential to the cortical surface at the target location.

Conventional neuronavigation systems typically use a plan-on-scalp or smooth volumetric brain model approach, in which users manually define target points on the scalp or a simplified 3D brain surface to construct the resulting stimulation trajectory (Ambrosini et al., 2018; Cline et al., 2025). This approach does not explicitly account for cortical geometry, as the three-dimensional brain model used for coil alignment is usually derived from a volumetric MRI reconstruction rather than a surface-based cortical topography. As a result, coil positioning relies heavily on subjective alignment without incorporating a subject-specific cortical topography. Figure 1c shows a comparison between the coil pose planned by commercial neuronavigation software and by the proposed topography-guided algorithm.

Once a coil pose is determined, a grid of targets can be defined depending on the shape of the cortical surface at different target points (Figure 1d). This strategy leverages the detailed geometry of the cortical surface, offering finer, more deterministic control over the planned coil poses. Due to the complex shape of the gyri and sulci on the cortical surface, the scalp is not always naturally parallel to the underlying cortical topography. As a result, when the contacting surface of the coil is aligned tangentially to the cortical surface, it may be angled in a way that causes collision with the scalp, preventing it from proper alignment parallel to the cortical surface (Figure 1e). In these cases, we use a slack solution to ensure the coil placement remains tangential to the scalp surface while being oriented as closely as possible to the target cortical location. Like the previous method, a target pose is placed on the cortical surface. Then, the closest point on the skin surface is found by applying Nearest Neighbor Search (NNS) on the surface mesh. The NNS approach locates the closest entry point on the skin by finding the point **s**^∗^∈ *S* with minimal Euclidean distance to the target:

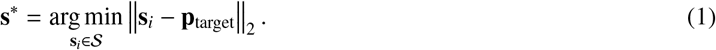

Here **p**_target_ denotes the planned target, and the scalp is represented as a discrete point set 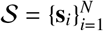 extracted from the reconstructed head mesh. Therefore, the final pose is established based on the projection of the desired cortical target point on the scalp, with a loose constraint induced by the curvature of the underlying brain. This approach aims to achieve accurate and reproducible TMS coil alignment compared to conventional methods, which depend on the visual alignment of the coil by the operator.

### 2.2. Refined Head and MRI Co-registration

Co-registration establishes a spatial relationship (transformation) between the MRI space and the physical world, which is necessary for accurate placement of the TMS coil at a designated target through the neuronavigation system. TMS co-registration commonly relies on paired-point strategy, which aligns a small set of anatomical landmarks on the subject’s head with their corresponding locations in the MRI, to solve the transformation between the MRI coordinate system {*M*} and the head reference frame {*H*}.

During this procedure, five anatomical landmarks were selected on the head at the nasal tip (pronasale), nasion, left and right tragus, and occipital protuberance. An infrared (IR)-tracked stylus was used to digitize the coordinates of these points, which is the process of marking their locations within the optical tracker coordinate system {*O*} and subsequently converting them to the MRI coordinate system {*M*}, while strictly preserving point-to-point correspondence. This step aligns two coordinate systems with a residual Cartesian distance that can fall below 6 mm (Figure 2).

**Figure 2.**
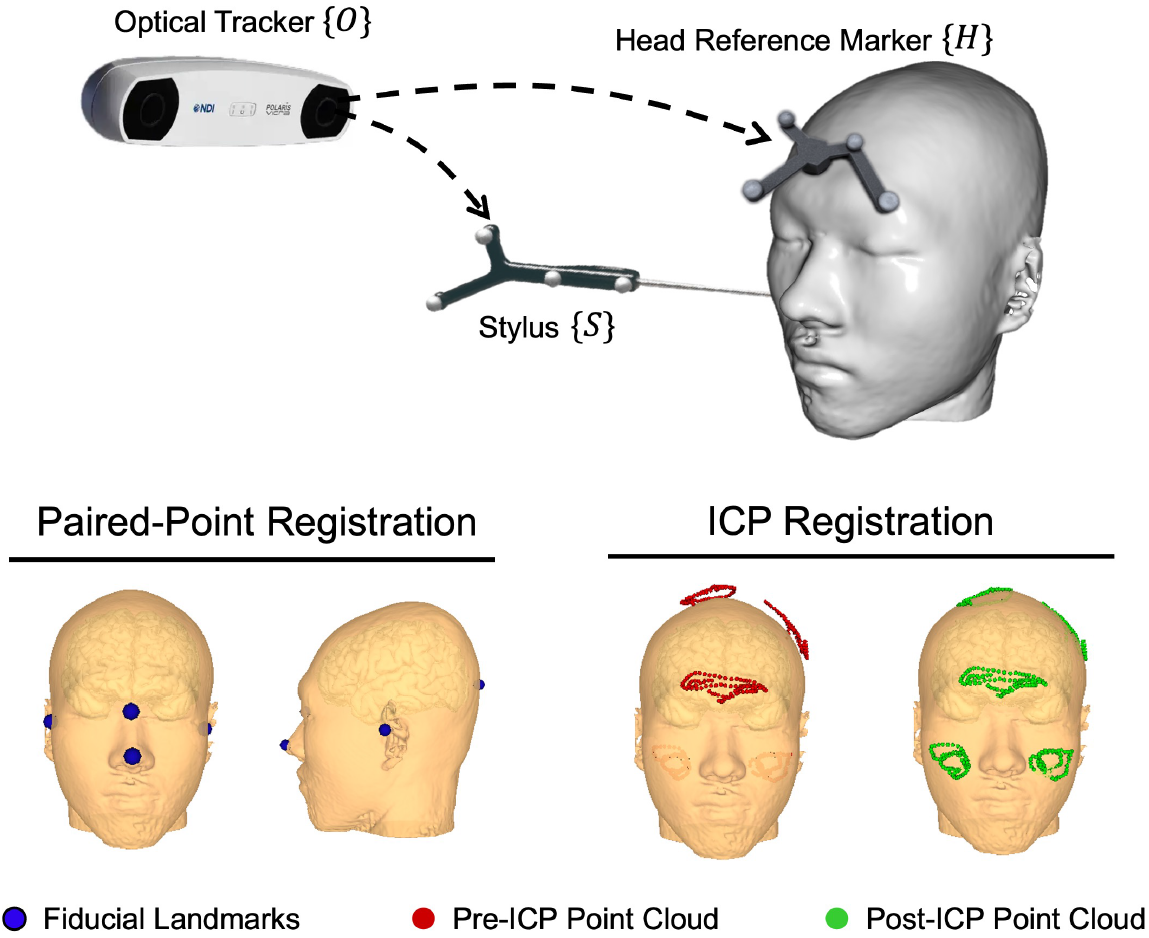
Co-registration procedure to obtain the transformation between the head and the corresponding MRI model. The procedure has two steps: 1) register anatomical landmarks; 2) collect a series of points on the head and use the Iterative Closest Point (ICP) algorithm to refine the transformation. To co-register anatomical landmarks or points on the head, a stylus {*S*} equipped with optical marker is used while a reference marker {*H*} tracks head position (dashed lines to optical tracker {*O*}). The landmarks used in the paired-point stage are highlighted as blue points on the MRI model. The red and green points represent the same set of sampled points in 3D space. Prior to registration, the misalignment between the sampled points and the reference surface is more pronounced, as illustrated by the red points, especially on top of the head. This misalignment also results in some points lying inside the skin surface, especially in the cheek region. After applying the ICP-based registration, this misalignment is corrected, resulting in improved alignment as shown by the green points.

The paired point registration is highly sensitive to noise and local registration errors because of using only a few landmarks. Surface-based registration improves robustness by refining the initial MRI-physical space alignment by sampling additional points with an IR-tracked stylus across bony regions, including the forehead, nose, zygomatic area, and vertex, to increase geometric constraints (Chiurillo et al., 2023; Nieminen et al., 2022; Noirhomme et al., 2004). This is especially relevant for neuronavigation guided by cortical topography, where the accuracy of stimulation depends on accurate registration of the head and MRI model.

We used the Iterative Closest Point (ICP) algorithm (Besl and McKay, 1992; Cline et al., 2025; Nieminen et al., 2022) to further refine the co-registration by reducing residual error between the sampled points on the head surface and the MRI model. Figure 2 illustrates the ICP-based refinement. The ICP algorithm computes an optimal rigid transformation by iteratively finding point correspondences and minimizing the point-to-point residuals between the sampled head surface and the corresponding MRI surface. Within each iteration, the ICP algorithm identifies nearest neighbors of the sampled points on the MRI surface and estimates the optimal rigid transformation using a least-squares method that minimizes the sum of squared distances between corresponding points.

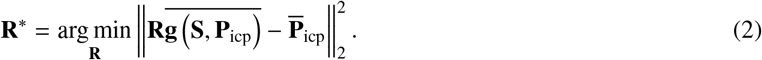

Here 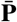 represents the centralized point set of **P**, i.e. 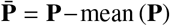, and **g** (**S, P**) denotes the operation of finding the nearest neighbor of point set **P** in a surface mesh set **S. P**_*icp*_ are point clouds collected on the head. Translation **t** can be solved after **R**^∗^ is found through:

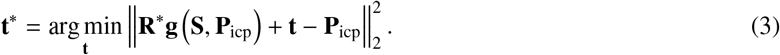

By iteratively applying these two steps, **R** and **t** will converge on the optimal solution **T**^∗^ = (**R**^∗^, **t**^∗^), which is the optimal transformation between {*M*} and {*H*}. For paired-point registration, the registration error was evaluated as Fiducial Registration Error (FRE), which is the Euclidean distance between corresponding anatomical landmarks on the head and the MRI model. For refined surface registration with cloud points, the registration error was evaluated across all collected points as the point-to-surface distance to the MRI model, quantified as the root mean square error:

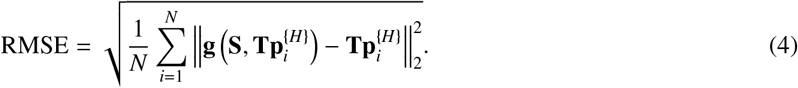

where *N* is the number of points.

### 2.3. Neuronavigation System

To implement TMS coil alignment based on cortical topography, we have developed a neuronavigation system that integrates the coil pose planning procedures described above with precise coil alignment using RA-TMS (Liu et al., 2024a). This neuronavigation system is built on 3D Slicer, an open-source platform for medical image processing and visualization. It consists of three core modules: the medical image planning module, robot control module, and target visualization module (Figure 3). The medical image planning module supports all pre-operative procedures, including surface-based co-registration, coil pose planning, and grid planning, during which coil trajectories are defined directly on the MRI. The intra-operative functions, including monitoring the robot pose, TMS coil pose, and co-registration accuracy, are managed within the target visualization module. The robot control module supports autonomous or manual robot execution. A middleware layer validates each operational step (for example, preventing robot motion until co-registration error falls within an acceptable range) and ensures real-time, reliable command dispatch between software and hardware (Liu et al., 2024a). The communication is established via Local Area Network for connecting the middleware, hardware, and user interface.

**Figure 3.**
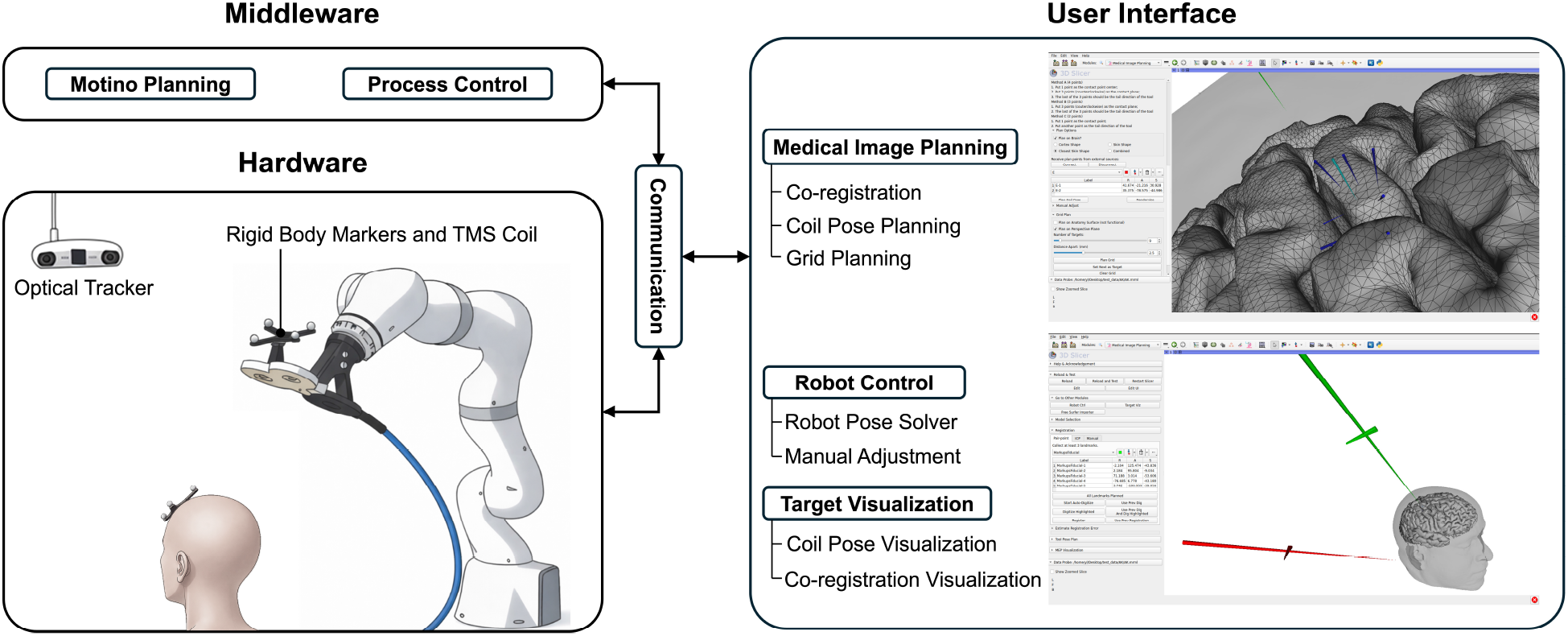
Architecture of the neuronavigation system used for applying TMS guided by cortical topography. The KUKA robot is used for the placement of the TMS coil. Rigid body markers are attached to the TMS coil and the subject’s head to enable tracking by the optical tracker within the neuronavigation system. The user interface is based on 3D Slicer and communicates with other hardware through a robot operating system (ROS) as middleware. The user interface supports preoperative planning and intraoperative control, and it provides necessary visualization for the operator to monitor, inspect, and review the entire procedure (Liu et al., 2024a,b). Target points are defined on the motor cortex, and both the planned coil pose (green cone) and the real-time pose adjusted by the robot (red cone) are displayed within the user interface. The cone represents the orientation of the TMS coil center, while the shorter axis perpendicular to the cone indicates the orientation of the coil handle.

## 3. Experiment

Here, we describe the experimental setup used to apply TMS at the primary motor cortex (M1) in human participants. We compared RA-TMS with manual coil positioning to determine their ability to support topography-guided TMS in practical use.

The neuronavigation system described above was used to control and visualize the procedures and to co-register the subject’s head with their MRI model. Magnetic pulses were generated using a MagStim Rapid2 stimulator and a 70 mm figure-of-eight TMS coil (Magstim Co., Whitland, UK). Two separate IR-reflective rigid body markers were attached to the TMS coil and the subject’s head. Each marker has a rigid body frame defined by four retro-reflective spheres and was localized by an Optical Tracking System (NDI Polaris Vicra, Northern Digital Inc., Canada). Delsys Electromyography (EMG) (Delsys Inc., MA, USA) was used to record the motor-evoked potential (MEP) at the left first dorsal interosseous (FDI), with one ground electrode over the left ulnar styloid process. The MEP was calculated based on the peak-to-peak amplitude of the EMG readings.

### Robot method

Our RA-TMS system uses a serial robotic manipulator (LBR iiwa 7 R800, KUKA, Germany) to carry the coil. The coil is mounted on the flange of the robot, and its spatial relationship with the robot is calibrated through a geometry-based method (Liu et al., 2024b). The robot is controlled by the neuronavigation software described above.

### Manual method

The same TMS setup was used here, but the coil attached to a mechanical arm was held manually by an expert operator. Real-time visualization of the coil pose and the subject’s head was provided within the neuronavigation software, allowing the operator to align the coil with the desired target orientation and maintain it manually throughout stimulation. This approach enabled a direct comparison between topography-guided RA-TMS and conventional manual TMS, in which the operator may maintain the coil near the target but not consistently preserve its precise position and orientation relative to the underlying cortical topography during stimulation.

#### 3.1. Experiment Protocol

The study protocol was reviewed and approved by the Institutional Review Board (IRB) of the Johns Hopkins University School of Medicine (Protocol number: NA_00032707; approval date: September 14th, 2013). Written informed consent was obtained from all participants before participation. All procedures were performed in accordance with institutional guidelines and applicable regulations, and participant privacy rights were observed. Participants underwent a high-resolution structural MRI (0.5 mm) scan to generate a three-dimensional brain model for neuronavigation. A reflective rigid-body marker was affixed to the forehead to enable optical tracking by the neuronavigation system. A total of 263 magnetic pulses were delivered in three subsets in separate sessions. During the experimental sessions, participants were seated in a fixed chair with their heads stabilized using a custom bite bar.

First, five targets, each 5 mm apart, were placed along the hand knob area within primary motor cortex (M1). At each target, the coil pose was computed based on the underlying cortical surface geometry, and five TMS pulses were delivered at 70% stimulator intensity. The target location where TMS elicited the maximal MEP, with at least three of five responses ≥0.05 mV, was selected (Grab et al., 2018; Reijonen et al., 2020). Next, a 3 × 3 square grid of targets spaced at 2.5 mm intervals was generated and centered on this location. Five TMS pulses were then delivered at each target within the grid, and the target with the highest mean MEP amplitude under the five pulses was defined as the hotspot location at M1.

At the hotspot location, the resting motor threshold (rMT) was determined as the minimum TMS intensity required to evoke MEPs ≥ 0.05 mV in the FDI muscle in at least 3 of 5 consecutive trials. (Rossi et al., 2021). We used a staircase method to determine rMT in which the TMS intensity was adjusted based on the following: if three or more MEPs exceeded 0.05 mV, the intensity was reduced initially by 5%, then 3%, and finally 1%. Conversely, if fewer than three MEPs exceeded 0.05 mV, the intensity was increased by 3% and then 1% to asymptotically reach the threshold intensity.

Robotic TMS guided by cortical topography was compared with manual TMS, in which cortical topography was not actively maintained during stimulation. Twenty consecutive TMS pulses were delivered at 105% rMT with interstimulus intervals of 10 seconds. To further assess physiological variability during stimulation guided by cortical topography, an additional 60 pulses were delivered using RA-TMS at 105% rMT with 10-second intervals. During all sessions, coil alignment error was quantified as the transformation between the planned and achieved coil poses. This error was decomposed into translational and rotational components.

## 4. Results

### 4.1. Refined Head and MRI Co-registration

Here, we report the errors for the paired point and refined registration methods. The FRE for the paired point method was 5.83 ± 1.04 mm (mean ± SEM), with variability primarily arising from the difficulty of consistently identifying corresponding anatomical landmarks on both the MRI and the subject’s head. Using a point cloud of 700 digitized surface points, the registration RMSE between the point cloud and the MRI surface was reduced to 1.86 ± 0.07 mm using the transformation obtained by the paired-point method. This improvement is attributed to the use of nearest-neighbor search for point correspondence, rather than manual landmark annotation. After using ICP, the registration error was further reduced to 1.07 ± 0.03 mm using the ICP method. Notably, 99.9% of points in the point cloud had their distance to the head surface less than 5 mm after ICP refinement (inliers), whereas only 81.8% points of the point cloud were under this threshold using the transformation obtained by the paired-point method.

### 4.2. Topography Guided RA-TMS vs. Manual TMS

To compare the manual and robotic methods, 20 consecutive TMS pulses at 105% of rMT were applied at the hotspot location at M1 (Figure 4a). There were significantly higher positional errors in the manual method compared to the robotic method (Figure 4b). The mean translational error for manual placement was 14.79 ± 8.03 mm (mean ± SEM), whereas the robotic placement achieved 5.22 ± 1.68 mm. Similarly, the rotational error decreased from 7.01^°^ ± 2.29^°^ in the manual method to 1.09^°^± 0.33^°^ in the robotic method (*p* < 0.001, paired t-test). In addition to improved alignment, the robotic method resulted in substantially larger MEPs, with amplitudes approximately one order of magnitude higher than manual TMS (0.27 ± 0.05 mV vs. 0.03 ± 0.01 mV, *p* < 0.001, paired t-test) (Figure 4b).

**Figure 4.**
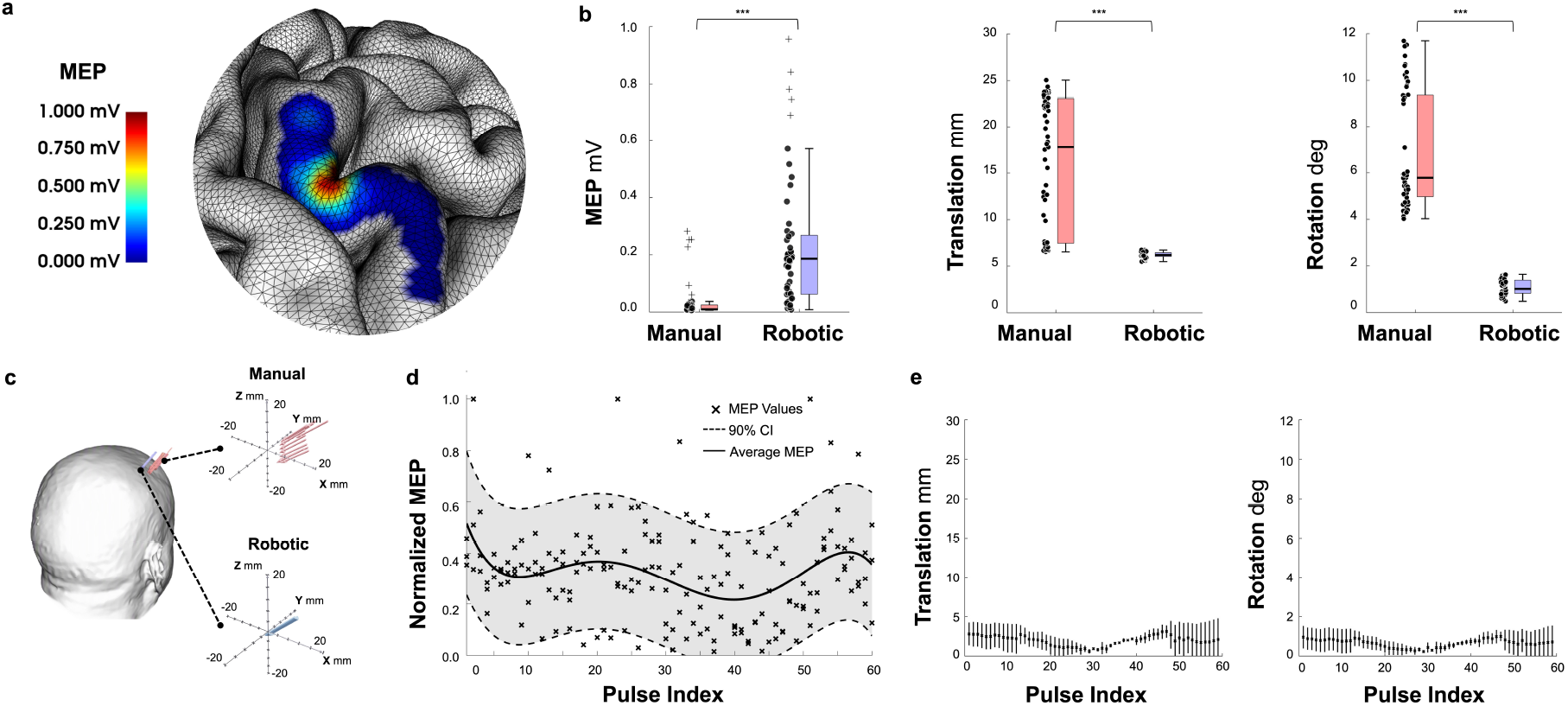
**a.** Example of hotspot localization at M1 shown as a cortical surface colormap derived from average MEP amplitudes across 13 target locations, defined by average MEP responses and visualized as a colormap on the cortical surface. The colormap is generated from MEP amplitudes at each target, with values decreasing radially within a 2.5 mm radius. Overlapping color values are averaged to produce a smooth, continuous distribution of MEP intensity across the cortical surface. **b**. Average MEP responses (left) at the M1 hotspot, along with coil alignment error during 20 consecutive TMS pulses in the manual (red) versus robotic (blue) methods, decomposed in translation (middle) and orientation (right). TMS coil alignment errors are decomposed into translation and rotational errors. (∗ ∗ ∗, *p* < 0.001). **c**. Coil pose distribution on the scalp and in 3D coordinate frame. The center of each plot represents the planned target, and the arrows indicate the coil pose at each pulse. Twenty consecutive pulses are shown for manual TMS (red) and RA-TMS (blue). Although scalp displacement was minimal with both methods, manual TMS showed greater orientational errors relative to the cortical target. **d-e**. MEP responses using RA-TMS with corresponding coil alignment error during a longer session with 60 consecutive pulses delivered at the M1 hotspot in the same subjects. Individual responses are normalized to maximum MEP amplitude (black crosses) and fitted using a polynomial basis function (black curve), with the 90% confidence interval (dotted lines). Alignment errors remained low throughout the longer stimulation session.

Figure 4d shows the response amplitude, and Figure 4e shows coil alignment error during a longer stimulation session with a train of 60 single pulses using RA-TMS. With low placement errors (translation: 1.96 ± 1.33 mm, orientation: 0.65^°^ ± 0.43^°^), the overall response pattern reflects physiological variability with larger MEP early on during the stimulation that decreased after several pulses and then recovered again despite the constant stimulation intensity and minimal placement error.

## 5. Discussion

In this study, we validated a fully integrated robot-assisted TMS system guided by subject-specific cortical topography. A central contribution of this work is the incorporation of detailed cortical geometry into coil pose planning. Unlike conventional neuronavigation systems that rely on simplified volumetric models (Cline et al., 2025), our approach explicitly accounts for the local cortical geometry by aligning the induced electric field perpendicular to a tangential plane at the target cortical location. This anatomically informed approach addresses a fundamental limitation of current TMS practice, where coil orientation is often approximated and may not reflect the true geometry of the underlying cortical gyri and sulci.

Leveraging a surface-based model that captures detailed cortical geometry enables deterministic and reproducible coil alignment tailored to focal cortical topography. This approach supports placement of individual or a grid of stimulation targets over the cortical surface. At each target location, coil position and orientation are derived directly from the cortical surface or from the nearest point on the scalp surface to the cortical target, depending on whether the cortex-restricted pose or the closest-scalp pose yields optimal placement. This strategy extends RA-TMS beyond improvements in mechanical stability toward anatomically grounded alignment of the TMS coil. This enhanced targeting approach substantially improved both the accuracy and consistency of stimulation outcomes, as demonstrated by MEP responses recorded at the motor cortex.

Robotic systems, first applied for TMS in early 2000s (Lancaster et al., 2004), have offered more stability and precision compared to manual coil placement, especially for long TMS sessions (Goetz et al., 2019). Much of the progress in robot-assisted TMS has largely focused on system design, workflow, calibration techniques, and clinical implementation (Grab et al., 2018; Matsuda et al., 2021, 2024b; Noccaro et al., 2021; Richter, 2013; Giuffre et al., 2021). Early implementations relied on industrial robots and were later augmented by optical tracking and force or torque sensing to improve safety and compensate for head motion (Matthäus et al., 2006; Pennimpede et al., 2013; Zorn et al., 2012; Richter et al., 2010; Matsuda et al., 2024b). Despite these advances, most existing RA-TMS systems implicitly assume that maintaining a fixed coil pose on the scalp is sufficient for reliable stimulation. Our approach here directly addressed this limitation by integrating robotic coil placement, planning, and alignment based on the underlying cortical topography.

Accurate implementation of TMS guided by cortical topography also depends critically on precise co-registration between the subject’s head and MRI space. Here, we used a surface-based refinement with ICP registration that significantly reduced spatial error compared to conventional paired-point methods (Ginhoux et al., 2013). By aligning dense point clouds rather than relying on manually selected anatomical landmarks, this approach reduces registration error. This level of accuracy is essential for ensuring that planned coil poses correspond closely to the intended cortical targets, particularly when using high-resolution surface models. Despite advances in neuronavigation, maintaining precise coil alignment during stimulation remains a major challenge with manual methods. Our results demonstrate that although manual TMS allowed the operator to maintain the coil over the cortical target, RA-TMS markedly improved the precision of cortical topography-guided stimulation by maintaining both the coil position and its orientation relative to the underlying cortical surface. Compared to manual TMS, RA-TMS reduced translational error by nearly 50% and rotational error by over 90%, while also significantly reducing variability across trials and subjects. The robotic coil alignment guided by cortical topography also elicited larger and more reliable MEPs compared to manual stimulation.

These findings highlight the importance of robotic stabilization in maintaining TMS coil alignment with cortical topography to achieve a more reliable assessment of intrinsic cortical excitability by improving coil alignment and enhancing cortical stimulation. Our results show that during repetitive stimulations, MEP amplitudes exhibit dynamic fluctuations with an initial decline followed by partial recovery, despite constant stimulation intensity and minimal placement error. This response pattern may reflect transient neuronal depolarization or short-term adaptation leading to reduced cortical excitability with repetitive TMS followed by recovery (Chen et al., 2019; Kiers et al., 1993; Modugno et al., 2001). Peripheral mechanisms, such as minor fluctuations in muscle excitability or electrode impedance, cannot be entirely excluded, although the 10 s inter-pulse interval and stable recording conditions make this explanation less likely (Darling et al., 2006).

Several directions remain for future development. Markerless tracking approaches (Matsuda et al., 2023) could simplify setup and further improve registration procedures, and advanced robotic motion compensation strategies (Richter et al., 2010) could improve coil alignment in response to any head motion during stimulation. Active force sensing can further improve contact safety and maintain stable coil engagement (Jaroonsorn et al., 2020; Lin et al., 2018). Combined with electroencephalography (EEG) measurements (Parmigiani et al., 2025) and electric field modeling (Gomez et al., 2021), the neuronavigation software would be able to optimize the coil pose, which not only guarantees spatial alignment but also facilitates maximum delivery. Ultimately, these components could be integrated into a fully automated RA-TMS system capable of autonomous targeting, stimulation, and adaptation.

In conclusion, this study demonstrates that integrating high-resolution cortical modeling with robotic coil positioning achieves superior alignment accuracy and significantly enhances stimulation outcomes. These findings support the importance of cortical geometry in optimizing brain stimulation and the adoption of robotic assistance in TMS applications for both research and clinical use.

## CRediT Author Statement

**Jiaming Zhang:** Methodology, Software, Validation, Investigation, Data Curation, Writing - Original Draft, Visualization. **Yihao Liu:** Conceptualization, Methodology, Software, Validation, Investigation, Writing - Review & Editing. **Jing Tian:** Conceptualization, Methodology, Resources, Writing - Review & Editing. **Dale Roberts:** Validation, Investigation **Mehran Armand:** Conceptualization, Writing - Review & Editing, Supervision. **Amir Kheradmand:** Conceptualization, Methodology, Resources, Writing - Review & Editing, Supervision, Funding acquisition.

## Acknowledgment

This work was supported by National Institute of Deafness and Other Communication Disorders (R01DC018815).

## Data availability

The data that support the findings of this study are available from the corresponding author upon reasonable request, subject to institutional and ethical restrictions.

